# Human Papillomavirus (HPV) self-testing among un- and under-screened Māori, Pasifika, and Asian women in Aotearoa New Zealand: a preference survey among responders and interviews with clinical-trial non-responders

**DOI:** 10.1101/2022.03.29.22273037

**Authors:** Susan M. Sherman, Karen Bartholomew, Naomi Brewer, Collette Bromhead, Sue Crengle, Chris Cunningham, Jeroen Douwes, Sunia Foliaki, Jane Grant, Anna Maxwell, Georgina McPherson, John D. Potter, Nina Scott, Helen Wihongi

## Abstract

**Aim:** Māori, Pasifika, and Asian women are less likely to attend cervical screening and Māori and Pasifika women are more likely to be diagnosed with later-stage cervical cancer than other women in Aotearoa New Zealand. This study – with under-screened women taking part in a randomised controlled trial comparing self-testing and standard screening – explored the acceptability of an HPV self-test kit and the preferred method for receiving it.

**Methods:** Māori, Pasifika, and Asian women (N=376) completed a postal questionnaire. Twenty-six women who had not accepted the trial invitation were interviewed to understand their reasons for non-participation.

**Results:** Most women found the self-test kit easy and convenient to use and reported that they did not find it painful, uncomfortable, or embarrassing. This was reflected in the preference for a self-test over a future smear test on the same grounds. Most women preferred to receive the kit by mail and take the test themselves, rather than having it done by a doctor or nurse. There was a range of preferences relating to how to return the kit. Phone calls with non-responders revealed that, although most had received the test kit, the reasons for not choosing to be involved included not wanting to, being too busy, or forgetting.

**Conclusions:** HPV self-testing was acceptable for Māori, Pasifika, and Asian women in Aotearoa New Zealand. HPV self-testing has considerable potential to reduce the inequities in the current screening programme and should be made available with appropriate delivery options as soon as possible.

## INTRODUCTION

Globally, cervical cancer is the fourth most common cancer affecting women, with around 570,000 diagnoses and 311,000 deaths in 2018^1^. In Aotearoa New Zealand, there were 190 diagnoses and 72 deaths in 2018; cervical cancer is the fourth most common cancer in women aged 15-44 years^2^. Māori women have a 1.7x higher incidence of this cancer (age-standardised rate of 9.4 cases per 100,000) than non-Māori (5.4 cases per 100,000) and the mortality rate is 2.7 times higher for Māori women (3.0 deaths per 100,000) than non-Māori (1.1 cases per 100,000)^3^. A review of cervical cancer incidence in 2013-2017^4^ showed that, for those women who were screened in the 6-84 months before diagnosis, 40% of Māori women and 53% of Pasifika^Footnote1^ women had a high-grade cervical cytology sample compared to 16% of European women, suggesting missed opportunities for prevention or earlier diagnosis. One of the key recommendations of the review was that resources be “provided to *improve access to screening* and treatment of cervical precancer for Māori women…. *Intervention strategies should take into consideration both the practical and cultural needs* of these women/wāhine.” (^4^p14, emphasis added).

High-risk human papillomavirus (HPV) types, primarily transmitted by sexual contact, are detected in more than 90% of cervical cancers^5^. Persistent infection with high-risk HPV can result in precancerous changes in cervical cells, which, if left untreated, can progress to cervical cancer. Currently, the National Cervical Screening Programme (NCSP) in Aotearoa New Zealand recommends cervical screening every three years for women aged 25-69, with cytology as the primary test^6^. HPV testing is an alternative primary screening test and many countries have already implemented this in their programmes.

Overall three-year screening coverage in Aotearoa New Zealand dropped from 72.2% in March 2019 to 70.2% in March 2021^7^ against a target of 80%. For Māori women, the reduction was from 64.6% to 61.2%; for Pasifika women it was 69.4% to 63.1%; and for Asian women it was 62.4% to 61.4%. The existing inequitable outcomes for Māori and Pasifika women are likely to worsen if these declines in screening continue. Moving to a system of primary HPV screening opens the possibility of offering women the option of HPV self-testing. This can be done in women’s homes or in a clinic setting and may address patient/client- and provider-related barriers to screening that have arisen due to structural and systemic biases.

A previous study that explored barriers to cervical screening for Māori women, as well as hypothetical acceptability of HPV self-testing^8^, found that the primary barriers to conventional, more invasive screening were whakamā/embarrassment^Footnote2^, lack of time/other commitments, and fear of discomfort or pain. In that study, 61.2% of women said they would prefer an HPV self-test to either clinician-taken HPV test or conventional screening and 73.3% said they would be likely or very likely to self-test if it were offered. A pilot study exploring acceptability found that 66% of under-screened Māori and Pasifika women preferred to self-test after trying a device^9^. Subsequently, two randomised controlled trials (RCTs) have been conducted to explore HPV self-testing with under- or never-screened women, both with a focus on Māori. In the first study, women in the intervention arm of the RCT were offered an HPV self-test, although they could opt instead for a clinician-taken sample or a smear test. The self-test could be done at home, in the clinic, or at a community centre. The control arm was usual care, so the women were offered a clinician-taken smear test. Of women in the self-test arm of the RCT, 59% were screened, compared to 22% of those in the usual-care arm, suggesting that an offer of self-testing could substantially increase screening uptake among Māori women^11^.

A second, larger, RCT conducted by our research team^9^ was preceded by a feasibility study^12^, which focused on stakeholder co-design and testing the cultural appropriateness of materials. Between 2018 and 2020, an open-label, three-arm, community-based, randomised, controlled trial in which un- or under-screened (≥5 years overdue) Māori, Pasifika, and Asian women from Auckland were invited for cervical screening was undertaken. The three trial arms were: usual care in which women were invited to attend a clinic for a standard smear sample; clinic-based self-testing in which women were invited to take a self-test at their usual general practice; and mail-out self-testing in which women were posted a kit and invited to take a self-test at home. This showed that, although screening uptake was lower than in the first RCT, self-testing uptake was again statistically significantly preferred over usual care, with highest participation in the mail-out self-test arm^13^.

We report here the findings from the acceptability survey that was given to women participating in the self-testing arms of the second RCT and nested sub-study in which some non-responding women were offered opportunistic self-sampling when they presented to the clinic for other reasons^9^, as well as the findings from a telephone survey conducted with non-responding women. The main aims of the survey and interviews were: to explore the acceptability of the HPV self-test^Footnote3^ kit; to determine what preferences women had for invitation, sample return, and follow-up methods; to establish whether the level of information in the participant material was appropriate and acceptable for Māori, Pasifika, and Asian women; and to determine whether further localisation or refinement was required.

## METHOD

### Design

We conducted a paper-based cross-sectional survey throughout the main trial and nested sub-study, which recruited women between June 2018 and May 2020. We also conducted telephone interviews with women who did not respond to the invitation to take part in the trial, to understand their reasons for non-participation.

### Participants

All Māori, Pasifika, and Asian women who were invited to take part in one of the two self-testing arms of the RCT^13^ were given a survey to complete about their experience of self-testing. The women were all aged 30-69 years, were resident in Waitematā or Auckland District Health Board (DHB) areas and had never been screened or were overdue (≥5 years) for cervical screening.

A random sample of women who were invited to the RCT but did not take part (non-responders) were contacted by telephone to take part in a telephone interview. The aim was to recruit equal numbers of women from each ethnicity and study group.

### Measures - survey

The survey items were localised and developed (based on feedback) from the Australian iPap study-responders post-test questionnaire^14^ with permission from the iPAP investigators and these, along with some additional questions, are detailed below.

#### Questions about the cervical screening test

All women were asked: about their experience of the self-test kit; whether the instructions were clear and easy to understand; whether they had watched the study video clips (the women were directed to a webpage with clips that had subtitles in several languages: Te Reo Māori, Tongan, Samoan, Korean, and Simplified Chinese); if so, which ones; and for any other comments.

Women who had previously had a smear test were asked questions comparing their experience of the self-test kit with their experience of a smear test.

All women were presented with a list of 19 possible reasons^9^ why they might not have had a smear test recently – or ever; they were asked to identify all the reasons that applied, to further identify their main reason, and to provide any additional reason(s).

Women were asked questions regarding how they would like to have a cervical screening test in the future. They were asked to identify their two main reasons for this choice from a list of six options, which varied depending on whether they would prefer to have a test taken by a doctor/nurse or to receive a self-test kit. If relevant, they were asked follow-up questions exploring preferences for the collection and return of the self-test kit. Finally, they were asked whether they would recommend the self-test kit to a friend or whānau/family member and whether they would be more likely to take part in regular screening if they were able to do the test themselves.

#### Socio-demographic questions

The final part of the survey was a series of socio-demographic questions regarding: highest level of schooling; household income; which generation of their family came to Aotearoa New Zealand; whether English is their first language; and whether they identify as Māori.

### Measures – interviews

A structured interview guide was used with non-responders to explore whether they had received the invitation to the trial and their reasons for not taking part as detailed in Table 1.

**Table 1.** Interview guide for non-responders to the clinical trial.

| Question | Example script | Action |
| --- | --- | --- |
| Did they receive the invitation? | *I'm calling from the Women's Health self-test study - did you receive the invitation we sent you?* | If no, brief explanation and offer to send kit.<br>Update address details |
| If yes, enquire into their response to the invitation for example:<br>Was the invitation clear?<br>Did they understand what they were being asked to do?<br>Did they decide not to participate? | *Did you get a chance to have a look at it?*<br>*How did you find the info we sent you?*<br>*Was it clear what you were being invited to do?*<br>*Was there a reason that you didn't do the self-test?* | Record any comments.<br>Offer follow-up call from nurse if appropriate. |
| If appropriate, find out what | *Do you have any suggestions?* | Record any comments. |
| would have made them participate. | *Is there anything we could have done differently?* |  |

### Ethics

The study was approved by the Aotearoa New Zealand Northern B Health and Disability Ethics Committee (HDEC) (reference: 17/NTB/120). The Aotearoa New Zealand Ministry of Health (including the National Kaitiaki Group - a group of Māori women established to monitor the safe use of Māori data within the NCSP) and the participating DHBs, primary health organisations, and clinics approved the use of data to identify and contact eligible women.

### Analysis

The survey data are summarised using descriptive statistics. We ran a series of Pearson’s Chi Square tests to compare reasons for not ever or not recently having had a smear test across ethnicity.

To analyse the open-ended responses to survey questions and the telephone interviews, we undertook content analysis using an emergent-coding approach, whereby codes were identified from the data rather than *a priori*^15^. The interview data consist of short accounts of the conversation documented by the Māori interviewer making the call.

Ethnicity data were collected and categorised in line with the HISO 10001:2017 Ethnicity Data Protocols, whereby respondents self-identify and responses are categorised according to Statistics NZ descriptors (https://www.health.govt.nz/publication/hiso-100012017-ethnicity-data-protocols). We used a prioritised output (if multiple ethnicities are identified, only one is included in the analyses), ordered Māori>Pasifika>Asian>Other as per the Ethnicity-data standards.

## RESULTS

The results reported below are from those women who responded to the invitation to take part in the study.

### Interviews with non-responders

Interviews with non-responders were also conducted. One hundred and twelve contact attempts were made, at different times of the day. In total, 26 women were interviewed, 16 of these were from the self-test in the clinic group (Māori N=4, Pasifika N=4, Asian N=6) and 12 were from the self-test at home group (Māori N=4, Pasifika N=4, Asian N=4). The content analysis of the notes made by the interviewer generated 25 unique codes. The codes along with a frequency count are presented in Supplementary Materials.

### Survey

In total, 376 women (75.7% of women who returned their test) completed the survey (mean age 46.5 years, SD=10.6, range 30 to 71). Participants’ characteristics are presented in Table 2.

**Table 2.** Characteristics of women who completed the survey.

| Demographic questions | Level | n (%) |
| --- | --- | --- |
| Ethnicity | Māori | 108 (28.7) |
|  | Pasifika | 105 (27.9) |
|  | Asian | 163 (43.4) |
| Which generation of family came to New Zealand? | Born in Aotearoa New Zealand | 132 (35.1) |
|  | Moved to New Zealand from another country | 174 (46.3) |
|  | Parents moved to New Zealand | 24 (6.4) |
|  | Grandparents moved to New Zealand | 8 (2.1) |
|  | Family moved to New Zealand before grandparents were born | 4 (1.1) |
|  | Prefer not to say | 8 (2.1) |
|  | Left blank | 26 (6.9) |
| English is first language | Yes | 183 (48.7) |
|  | No | 162 (43.1) |
|  | Left blank | 31 (8.2) |
| Highest level of schooling | Primary school | 9 (2.4) |
|  | Secondary school (college) | 105 (27.9) |
|  | Technical or trade school diploma | 56 (14.9) |
|  | Undergraduate university degree | 89 (23.7) |
|  | Postgraduate university degree | 83 (22.1) |
|  | None | 8 (2.1) |
|  | Left blank | 26 (6.9) |
| Household's approximate gross (before tax, levies, etc) annual income | \$1 - \$20,000 | 27 (7.2) |
| | \$20,001 - \$50,000 | 81 (21.5) |
| | \$50,001 - \$70,000 | 48 (12.8) |
| | \$70,001 - \$100,000 | 59 (15.7) |
| | \$100,001 - \$150,000 | 32 (8.5) |
| | \$150,001 or more | 26 (6.9) |
|  | Prefer not to say | 66 (17.6) |
|  | Left blank | 37 (9.8) |

### Using the self-test kit

Women were asked to indicate the extent to which they agreed with a series of statements about the HPV self-test kit. The responses are presented in Table 3.

**Table 3.**
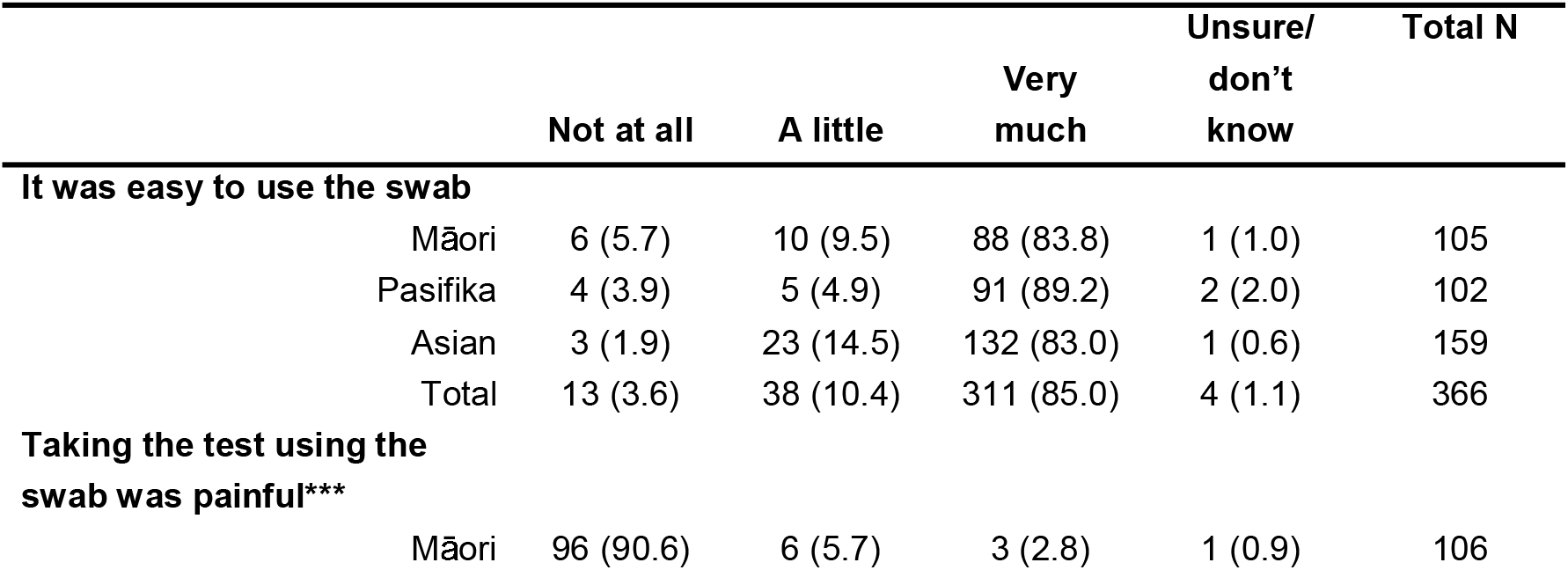
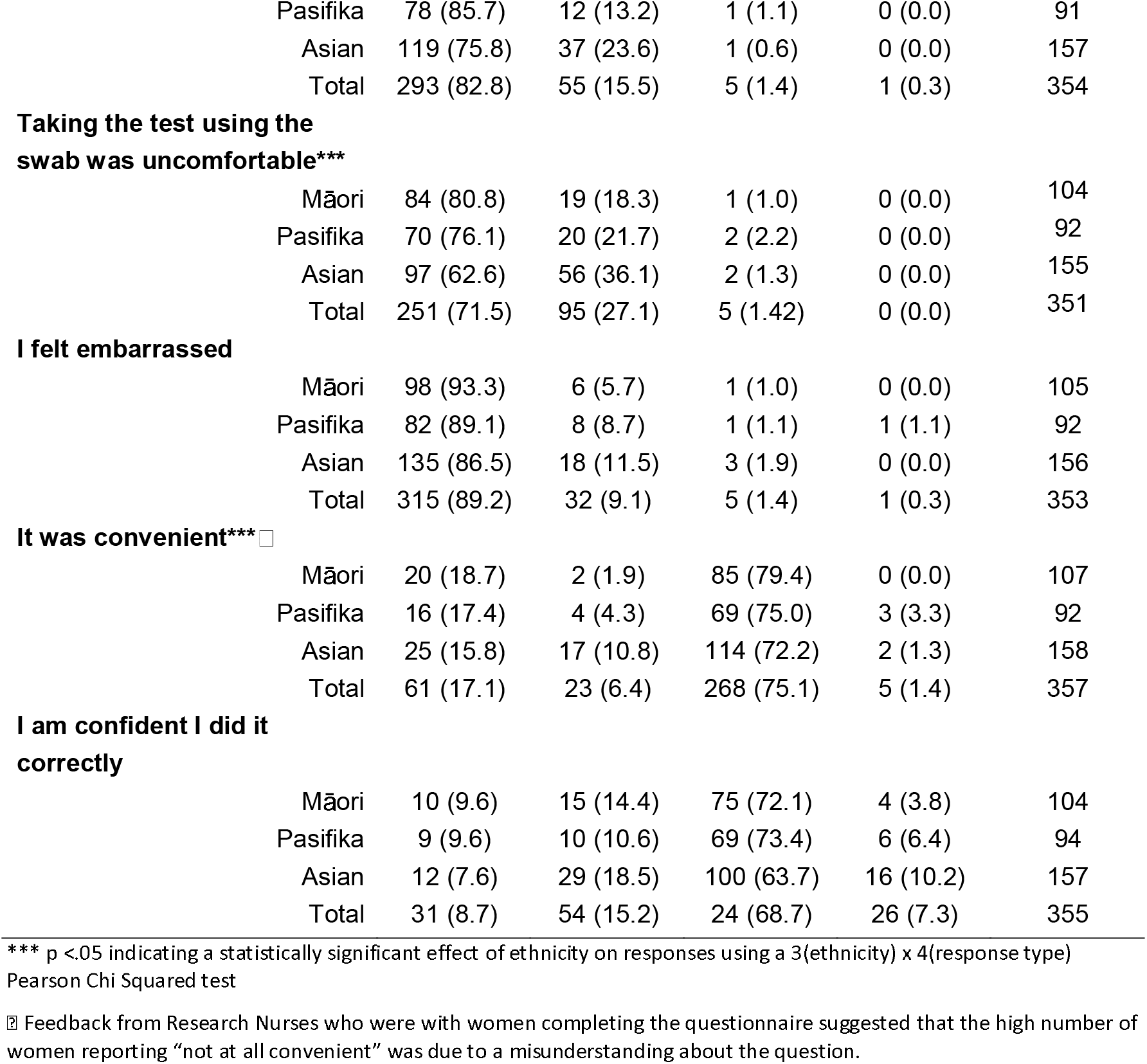
Women’s responses to statements about using the self-test kit (% in brackets)

|  |  | Not at all | A little | Very much | Unsure/<br>don't know | Total N |
| --- | --- | --- | --- | --- | --- | --- |
| **It was easy to use the swab** |  |  |  |  |  |  |
|  | Māori | 6 (5.7) | 10 (9.5) | 88 (83.8) | 1 (1.0) | 105 |
|  | Pasifika | 4 (3.9) | 5 (4.9) | 91 (89.2) | 2 (2.0) | 102 |
|  | Asian | 3 (1.9) | 23 (14.5) | 132 (83.0) | 1 (0.6) | 159 |
|  | Total | 13 (3.6) | 38 (10.4) | 311 (85.0) | 4 (1.1) | 366 |
| **Taking the test using the swab was painful\*\*\*** |  |  |  |  |  |  |
|  | Māori | 96 (90.6) | 6 (5.7) | 3 (2.8) | 1 (0.9) | 106 |

|  |  |  |  |  |  |
| --- | --- | --- | --- | --- | --- |
| Pasifika | 78 (85.7) | 12 (13.2) | 1 (1.1) | 0 (0.0) | 91 |
| Asian | 119 (75.8) | 37 (23.6) | 1 (0.6) | 0 (0.0) | 157 |
| Total | 293 (82.8) | 55 (15.5) | 5 (1.4) | 1 (0.3) | 354 |
| **Taking the test using the swab was uncomfortable\*\*\*** |  |  |  |  |  |
| Māori | 84 (80.8) | 19 (18.3) | 1 (1.0) | 0 (0.0) | 104 |
| Pasifika | 70 (76.1) | 20 (21.7) | 2 (2.2) | 0 (0.0) | 92 |
| Asian | 97 (62.6) | 56 (36.1) | 2 (1.3) | 0 (0.0) | 155 |
| Total | 251 (71.5) | 95 (27.1) | 5 (1.42) | 0 (0.0) | 351 |
| **I felt embarrassed** |  |  |  |  |  |
| Māori | 98 (93.3) | 6 (5.7) | 1 (1.0) | 0 (0.0) | 105 |
| Pasifika | 82 (89.1) | 8 (8.7) | 1 (1.1) | 1 (1.1) | 92 |
| Asian | 135 (86.5) | 18 (11.5) | 3 (1.9) | 0 (0.0) | 156 |
| Total | 315 (89.2) | 32 (9.1) | 5 (1.4) | 1 (0.3) | 353 |
| **It was convenient\*\*\*☐** |  |  |  |  |  |
| Māori | 20 (18.7) | 2 (1.9) | 85 (79.4) | 0 (0.0) | 107 |
| Pasifika | 16 (17.4) | 4 (4.3) | 69 (75.0) | 3 (3.3) | 92 |
| Asian | 25 (15.8) | 17 (10.8) | 114 (72.2) | 2 (1.3) | 158 |
| Total | 61 (17.1) | 23 (6.4) | 268 (75.1) | 5 (1.4) | 357 |
| **I am confident I did it correctly** |  |  |  |  |  |
| Māori | 10 (9.6) | 15 (14.4) | 75 (72.1) | 4 (3.8) | 104 |
| Pasifika | 9 (9.6) | 10 (10.6) | 69 (73.4) | 6 (6.4) | 94 |
| Asian | 12 (7.6) | 29 (18.5) | 100 (63.7) | 16 (10.2) | 157 |
| Total | 31 (8.7) | 54 (15.2) | 24 (68.7) | 26 (7.3) | 355 |

\*\*\* p <0.05 indicating a statistically significant effect of ethnicity on responses using a 3(ethnicity) x 4(response type) Pearson Chi Squared test
☒ Feedback from Research Nurses who were with women completing the questionnaire suggested that the high number of women reporting “not at all convenient” was due to a misunderstanding about the question.

In an additional question, of the 354 women who answered the question, 99.4% (N=352) said the self-test kit instructions were clear and easy to understand, with only 2 women indicating that they were not. Of the 358 women who answered the question, 78.5% (N=281) said they had not watched the video-clips while 21.5% (N=77) said they had. The videos watched were: About the HPV self-test study (N=59); How to take part in the study (N=50); How to do the test (N=58); Getting your test results (N=44); and About Cervical Screening And Your Rights (N=44). Sixty-seven women indicated that the video clips were helpful with just five women saying they were not.

Forty-two women provided responses other than N/A or ‘I didn’t watch them’ to the open-ended question asking for comments about the video clips (see Supplementary Materials).

### Comparison of self-test kit with previous smear test

Women who had previously had a smear test were asked to compare this with the self-test kit and their responses are presented in Table 4.

**Table 4:** Comparison of smear and self-test kit.

|  |  | Self-test | Smear test | No<br>difference | Unsure/don't<br>know | Total N |
| --- | --- | --- | --- | --- | --- | --- |
| **Easier** |  |  |  |  |  |  |
|  | Māori | 81 (92.0) | 2 (2.3) | 4 (4.5) | 1 (1.1) | 88 |
|  | Pasifika | 57 (80.3) | 6 (8.5) | 4 (5.6) | 4 (5.6) | 71 |
|  | Asian | 70 (79.5) | 6 (6.8) | 7 (8.0) | 5 (5.7) | 88 |
|  | Total | 208 (84.2) | 14 (5.7) | 15 (6.1) | 10 (4.0) | 247 |
| **More convenient** |  |  |  |  |  |  |
|  | Māori | 83 (96.5) | 1 (1.2) | 1 (1.2) | 1 (1.2) | 86 |
|  | Pasifika | 56 (88.9) | 4 (6.3) | 1 (1.6) | 2 (3.2) | 63 |
|  | Asian | 75 (89.3) | 3 (3.6) | 3 (3.6) | 3 (3.6) | 84 |
|  | Total | 214 (91.8) | 8 (3.4) | 5 (2.1) | 6 (2.6) | 233 |
| **Less embarrassing\*\*\*** |  |  |  |  |  |  |
|  | Māori | 81 (90.0) | 2 (2.2) | 6 (6.7) | 1 (1.1) | 90 |
|  | Pasifika | 57 (90.5) | 4 (6.3) | 2 (3.2) | 0 (0.0) | 63 |
|  | Asian | 69 (84.1) | 0 (0.0) | 9 (11.0) | 4 (4.9) | 82 |
|  | Total | 207 (88.1) | 6 (2.6) | 17 (7.2) | 5 (2.1) | 235 |
| **Less uncomfortable** |  |  |  |  |  |  |
|  | Māori | 79 (90.8) | 3 (3.4) | 4 (4.6) | 1 (1.1) | 87 |
|  | Pasifika | 55 (88.7) | 5 (8.1) | 2 (3.2) | 0 (0.0) | 62 |
|  | Asian | 67 (77.0) | 6 (6.9) | 10 (11.5) | 4 (4.6) | 87 |
|  | Total | 201 (85.2) | 14 (5.9) | 16 (6.8) | 5 (2.1) | 236 |
| **More accurate\*\*\*** |  |  |  |  |  |  |
|  | Māori | 15 (18.5) | 4 (4.9) | 5 (6.2) | 57 (70.4) | 81 |
|  | Pasifika | 22 (38.6) | 8 (14.0) | 1 (1.8) | 26 (45.6) | 57 |
|  | Asian | 11 (13.6) | 5 (6.2) | 5 (6.2) | 60 (74.1) | 81 |
|  | Total | 48 (21.9) | 17 (7.8) | 11 (5.0) | 143 (65.3) | 219 |

\*\*\* p < .05 indicating a statistically significant effect of ethnicity on responses using a 3(ethnicity) x 4(response type) Pearson Chi Squared test.

### Barriers to smear test

The number and percentage of reasons for not ever or not recently having had a smear test are presented in Table 5.

**Table 5.** Reasons for not ever or not recently having had a smear test (multiple selections were possible).

| Question | Māori | Pasifika | Asian | N (%)* |
| --- | --- | --- | --- | --- |
| A test from a nurse or doctor is embarrassing<sup>§</sup> | 60 (55.6) | 47 (44.8) | 57 (35.0) | 164 (43.6)*** |
| A smear test from a nurse or doctor is too painful or uncomfortable<sup>§</sup> | 40 (37.0) | 35 (33.3) | 47 (28.8) | 122 (32.4)*** |
| It is hard to find the time to have a test<sup>§</sup> | 33 (30.6) | 37 (35.2) | 49 (30.1) | 119 (31.6)*** |
| I don't think I need a test | 17 (15.7) | 19 (18.1) | 41 (25.2) | 77 (20.5) |
| I don't feel comfortable asking for a test from my nurse or doctor | 29 (26.9) | 21 (20.0) | 24 (14.7) | 74 (19.7) |
| I don't know if or when I should have a | 13 (12.0) | 21 (20.0) | 30 (18.4) | 64 (17.0) |
| test |  |  |  |  |
| I have had a bad experience in the past having a test | 29 (26.9) | 17 (16.2) | 15 (9.2) | 61 (16.2) |
| I am not having sex | 14 (13.0) | 16 (15.2) | 30 (18.4) | 60 (16.0) |
| It is hard to find or get an appointment with the right nurse or doctor | 9 (8.3) | 11 (10.5) | 8 (4.9) | 28 (7.4) |
| It is hard to travel to an appointment | 5 (4.6) | 6 (5.7) | 15 (9.2) | 26 (6.9) |
| It is too expensive to have a test | 7 (6.5) | 4 (3.8) | 9 (5.5) | 20 (5.3) |
| I have never had sex | 5 (4.6) | 3 (2.9) | 11 (6.7) | 19 (5.1) |
| I have not received a reminder letter to have a test | 5 (4.6) | 3 (2.9) | 11 (6.7) | 19 (5.1) |
| It is hard to find a nurse or doctor of the right sex | 10 (9.3) | 3 (2.9) | 6 (3.7) | 19 (5.1) |
| My nurse or doctor has not suggested a test | 2 (1.9) | 1 (1.0) | 10 (6.1) | 13 (3.5) |
| It is hard to find a nurse or doctor who speaks my language | 0 (0.0) | 2 (3.1) | 5 (1.9) | 7 (1.9) |
| I have had a hysterectomy | 3 (2.8) | 1 (1.0) | 3 (1.8) | 7 (1.9) |
| It is hard to find a nurse or doctor of the right ethnicity | 1 (0.9) | 3 (2.9) | 2 (1.2) | 6 (1.6) |
| I don't think test results are accurate enough | 0 (0.0) | 4 (3.8) | 1 (0.6) | 5 (1.3) |
| Rather not say | 3 (2.8) | 4 (3.8) | 2 (1.2) | 9 (2.4) |

¥ percentage is of all 376 participants, but multiple responses were allowed and included
\*\*\* p <.05 indicating a statistically significant effect of ethnicity on responses using a 3(ethnicity) x 2(selected reason or not) Pearson Chi Squared test
§ The top three reasons were also the most common responses by the 176 women who provided a main reason for lack of attendance from a list of options

Forty-one women provided a free-text self-reported reason for not having had a smear test (see Supplementary Materials).

### Future screening preferences

Women were asked to identify whether they would prefer to have a doctor or nurse do the test in the future or whether they would prefer to do it themselves and then to identify the two main reasons for choosing that answer. In total: 18 (4.8%) women indicated that they would prefer a doctor or nurse to take the test; 287 (76.3%) indicated that they would prefer to take their own test at home; 63 (16.8%) indicated that they would prefer to take their own test at a medical clinic; 3 (0.8%) said they did not intend to do a test again; and 7 (1.9%) women did not know what they would prefer. 18 women left the question blank. The reasons for their preferences are presented in Table 6 below.

**Table 6.** Reasons for preferring future tests to be administered by a doctor/nurse or self-administered (women were invited to select two reasons).

| Reasons for preferring a doctor or nurse to take the test | Māori (%) | Pasifika (%) | Asian (%) | N |
| --- | --- | --- | --- | --- |
| The test is accurate | 5 (4.6) | 7 (6.7) | 18 (11.0) | 30 |
| I can ask the nurse or doctor about something else*** | 1 (0.9) | 3 (2.9) | 13 (8.0) | 17 |
| The test is simple to do | 4 (3.7) | 5 (4.8) | 7 (4.3) | 16 |
| The test may find other problems | 3 (2.8) | 6 (5.7) | 7 (4.3) | 16 |
| The test is convenient | 2 (1.9) | 2 (1.9) | 6 (3.7) | 10 |
| The test is less embarrassing | 4 (3.7) | 1 (1.0) | 1 (0.6) | 6 |
| **Reasons for preferring to take the test yourself** |  |  |  |  |
| The test is less embarrassing | 61 (56.5) | 56 (53.3) | 75 (46.6) | 193 |
| The test is simple to do | 60 (55.6) | 52 (49.5) | 77 (47.2) | 189 |
| I do not need an appointment with a nurse or doctor to do the test | 54 (50.0) | 43 (41.0) | 87 (53.4) | 184 |
| The test does not require the use of instruments (e.g., speculum)*** | 48 (44.4) | 29 (27.6) | 47 (28.8) | 124 |
| The test is free | 25 (23.1) | 28 (26.7) | 48 (29.4) | 101 |
| The test is accurate | 4 (3.7) | 6 (5.7) | 4 (2.5) | 14 |
| *** p < .05 indicating a statistically significant effect of ethnicity on responses using a 3(ethnicity) x 2(selected reason or not) Pearson Chi Squared test |  |  |  |  |

Women’s preferences for receiving and returning the self-test kit if they used it in the future are presented in Table 7. If the self-test kit were to be mailed, 67.0% (N=252) said they would prefer the kit to be automatically sent the next time they were due for their smear test, whereas 23.4% (N=88) would prefer to receive a letter or call first; 2.1% (N=8) said they would prefer to order the kit online from a health professional. 28 women did not answer the question.

**Table 7.** Women’s preferences for receiving and returning the self-test kit if they were going to use it in the future.

| Preference | Māori (%) | Pasifika (%) | Asian (%) | N<sup>¥</sup> |
| --- | --- | --- | --- | --- |
| Receive self-test kit by mail*** | 83 (76.9) | 64 (61.0) | 124 (76.1) | 271 |
| Collect from clinic to do at clinic*** | 13 (12.0) | 20 (19.0) | 13 (8.0) | 46 |
| Collect from clinic to do at home | 11 (10.2) | 13 (12.4) | 13 (8.0) | 37 |
| Collect from pharmacy to do at home | 6 (5.6) | 4 (3.8) | 5 (3.1) | 15 |
| Collect from community laboratory to do at home | 3 (2.8) | 1 (1.0) | 2 (1.2) | 6 |
| Return kit by courier*** | 49 (45.4) | 28 (26.7) | 74 (45.4) | 151 |
| Return kit to a clinic (such as family doctor) | 30 (27.8) | 39 (37.1) | 49 (30.1) | 118 |
| Return kit to a community laboratory | 22 (20.4) | 20 (19.0) | 28 (17.2) | 70 |
| Return kit to a pharmacy | 11 (10.2) | 6 (5.7) | 11 (6.7) | 28 |

¥NB Some women selected more than 1 response, and some left the question blank, all responses were included
\*\*\* ps < .05 indicating a statistically significant effect of ethnicity on responses using a 3(ethnicity) x 2(selected reason or not) Pearson Chi Squared test

## DISCUSSION

The three main reasons Māori, Pasifika, and Asian women in our study provided for not ever or not recently having a smear test were embarrassment, pain or discomfort, and time. These findings are consistent with previous research,^8,16^ which found that the primary barriers to conventional screening for Māori women were whakamā/embarrassment, lack of time/other commitments, and fear of discomfort or pain. By contrast, most women in our study found the self-test kit to be easy and convenient to use and reported that they did not find it painful, uncomfortable, or embarrassing. This was also reflected in the preference for a self-test over a future smear test on the grounds of ease, convenience, comfort, and reduced embarrassment by most women who had previously had a smear test.

The only area of uncertainty around the self-test kit was related to the perceived accuracy of the self-test. A substantial minority (24%) were not confident or not sure that they had performed the test correctly. Furthermore, more than half the women (65%) who had previously had a smear test indicated that they were unsure whether the self-test kit or smear test was more accurate. In a focused literature review exploring acceptability, feasibility, and uptake of HPV self-testing among so-called hard-to-reach women across the world between 1997 and 2015, a key barrier to self-testing reported was a concern about sampling accuracy, including performing the procedure correctly^17^. This barrier was reiterated in a more recent scoping review of research exploring HPV self-testing in Indigenous communities^18^. The provision of more information addressing these areas may allay women’s concerns. Most women in our study (76%) indicated that they would prefer to take a self-test at home or in a clinic (17%) rather than having a clinician do the test (5%) and the main reasons for this were reduced embarrassment, the simplicity of the test, and not needing an appointment.

It is important to note that some of the findings differed by ethnicity. For example, although the three main reasons for not ever or not recently having had a smear test were the same for each group, the percentage of women from each group who reported these differed. For example, more Māori women than Pasifika women and more Pasifika women than Asian women reported embarrassment or pain or discomfort as a reason, while more Asian women than Māori or Pasifika women reported time as a reason. These and the other differences should be used to inform the design of information and educational materials to support HPV primary screening and self-testing which target at-risk groups.

The findings highlight areas where more information may be useful to increase uptake. For example, most women in our survey did not know whether the self-test or smear test was more accurate, 20% did not think they needed a test and, relatedly, 16% reported not having the test because they were not having sex. Our findings also give some clear insights into women’s preferences, which should be used to inform the implementation of HPV primary screening and self-testing. Most women (72%) would prefer to receive a self-test kit mailed to their home, although some women would prefer to collect it from a clinic to do at the clinic (12%) or at home (10%). Importantly, if the test were to be mailed to their home, although most women (67%) would be happy for this to happen automatically, a sizable minority (23%) would prefer to receive a letter or call first. In relation to returning the kit, findings were more mixed. Some women would prefer to return the test by courier, some would prefer to return it to a clinic, and others to a pharmacy or community laboratory. A range of options would provide the best rate of return, which is consistent with research that showed that a community laboratory drop-off alternative to postal return increased participation in New Zealand’s Bowel Screening Pilot^19^. As HPV self-testing is introduced, it will be important that these preferences are taken into account to maximise uptake of self-testing. A new purpose-built NCSP population register should allow recording of preferences for invitation and recall.

Maximising access to self-testing can also be informed by our phone calls with non-responders, which revealed that, although most had received the test kit, there were a variety of reasons for not doing the test, including not wanting to do it, being too busy, or forgetting (see Supplementary Material). Follow-up phone calls were very time consuming, so further research could investigate ways to maximise uptake of unsolicited self-testing kits, possibly with the use of advance notification that the kit was being sent and follow-up reminder letters, texts, or calls.

The aforementioned scoping review of research exploring HPV self-testing in Indigenous communities covered the years 1993 to 2018 and just 19 studies, including grey literature, met the criteria^18^; however, none of these were from Aotearoa New Zealand. The study reported here builds on previous research^9,16^, as well as our own feasibility study^12^ to provide the first research that explores the preferences around HPV self-testing of Māori, Pasifika, and Asian women who have actually used a self-testing kit. This research is timely for three reasons. First, the Aotearoa New Zealand Government has recently announced that they will invest “up to $53 million to … implement a new human papillomavirus (HPV) test” in 2023^20^. The screening programme will introduce the option of either a self-test or clinician-taken sample to all women, which means that it is crucial that culturally relevant research and Māori, Pasifika and Asian expertise and knowledge is used to inform the design and roll-out of the new programme. Second, HPV self-testing has recently been announced as an option for participants in the Australian National Screening Programme from July 2022^21^. Third, a recent editorial^22^ further suggests that the negative impact of the COVID-19 pandemic on cervical screening uptake may prove to be a tipping point for the wider introduction of self-testing. It is likely that Indigenous groups in other countries can benefit from these Aotearoa New Zealand findings.

To achieve elimination of cervical cancer as characterised by the WHO (4 cases per 100,000 women^23^), incidence must be reduced by 63% for Māori women^24^. This needs to be achieved by a concerted effort across primary care through HPV vaccination, cervical screening, diagnosis, and treatment/follow-up pathways. Our findings indicate that HPV self-testing would offer an acceptable and culturally appropriate addition to the screening programme in Aotearoa New Zealand for Māori, Pasifika, and Asian women – particularly if the preferences identified in our research are implemented – and thus has considerable potential to reduce inequities in access to screening and to save lives. Now that the Government has committed to rolling out HPV primary testing inclusive of self-testing, equity-focused approaches are required to ensure that benefits address the current and longstanding inequities in access to cervical screening and cervical cancer outcomes in Aotearoa New Zealand.

## Data Availability

As a result of ethics requirements and issues around data sovereignty associated with Indigenous people, at present we are unable to share data.

## Acknowledgements

We thank the staff of the participating Primary Health Organisations and GP clinics, and all of the women who took part in our study. We thank Marion Saville (Victoria Cytology Service) and Susan Reid (Health Literacy NZ) for assistance with the design of the study materials, and Mellissa Murray for assistance with interviews, project administration and coordination. Thanks are also due to the cultural advisory organisations: The Asian Network Inc (TANI), the Chinese New Settlers Trust and the Fono, and individual advisors: Samantha Bennett, Raj Kumar, Leani Sandford, and Sulu Samu. Thanks also to He Kamaka Waiora (Māori research services) and Waitematā DHB Asian Health Services who were available to take queries from participating women. We would also like to acknowledge that screening data for the study was obtained from the NCSP Register, with approval. The National Kaitiaki Group granted approval for the use of Māori women’s data from the register and reviewed the study manuscript.

## TRANSPARENCY DECLARATION

The authors affirm that the manuscript is an honest, accurate, and transparent account of the study being reported; that no important aspects of the study have been omitted; and that any discrepancies from the study as originally planned have been explained.

## CONFLICT OF INTEREST STATEMENT

N Brewer reports grants from Janssen-Cilag Pty Limited, outside the submitted work. C Bromhead has previously received educational funding to attend conferences from Roche Diagnostics New Zealand. S Crengle reports personal fees from Board member WellSouth Primary Health Network, personal fees from General Practitioner, Invercargill Medical Centre, personal fees from Board member, Royal NZ College of General Practitioners, outside the submitted work. H Wihongi reports grants and other from Waitematā District Health Board, outside the submitted work. She is also a member and convener of the National Kaitiaki Group. The other authors have no conflicts of interest to declare.

### CRediT roles

Conceptualization; KB, NB, CB, JD, AM, JP

Data curation; NB

Formal analysis; SS

Funding acquisition; KB, NB, CB, CC, JD, SF, GM, JP, HW

Investigation; All authors

Methodology; KB, NB, CB, SC, CC, JD, SF, AM, JP, NS, HW

Project administration; KB, NB, CB, JD, AM, JP

Resources; JD, JP

Supervision; KB, NB, CB, JD, AM, JP

Roles/Writing - original draft; SS Writing - review & editing; All authors

## Supplementary Materials

Additional data referenced in the text is presented here with relevant section headings from the main text.

### Codes generated by content analysis of the interviews with non-responders

Codes generated by content analysis of the interviews with non-responders.
| Code name | Number of comments per code | Indicative quotes or comments by interviewer |
| --- | --- | --- |
| Received pre-invite or kit | 19 | Received the invitation, forgot to complete her test. Found the information very helpful and clear. The mobile number we had was invalid, so she didn't know about the opportunistic invitation [for the sub-study], however, wanted a kit to be sent out if possible. [Asian clinic] |
| Wanted a kit to be sent | 7 | Never received the kit. 'Sounds good' if she doesn't have to 'have someone go down there'. Would like to have another look if not too late. Address updated and new kit sent out. [Pasifika home] |
| Information was good/liked the idea | 6 | The information was very helpful but with work and family had not got around to it yet. [Asian clinic] |
| Does not want to do the test | 6 | Received all information and texts. She did not do the test as she does not want to do it. When I asked if there was anything that would have encouraged her to, she said no she just did not want to do it. [Pasifika home] |
| Threw kit away/lost it | 5 | 1. "I have limited movement in my arms and neck ... I was a bit angry when I got the invite as how the hell am I going to do that? So I threw it away and didn't bother with it again" [Māori home]<br>When she got home she thought she had missed it so just threw it away. "I wasn't in NZ when the information was sent - I thought I had missed out. It would have been better if you had emailed me the information" [Pasifika home] |
| No invite received | 4 | Never received the information. Not really interested (not her thing) will just have a smear done next time. [Māori home] |
| Too busy to do the test | 4 | Received the invitation but has been busy with work and home. [Asian clinic] |
| Code name | Number of | Indicative quotes or comments by |

|  | comments per code | interviewer |
| --- | --- | --- |
| Forgot to do the test | 3 | Went into clinic and the nurse explained everything to her, she said it looked easy and simple. She took a kit home to do the test and life got busy and she forgot. However, because I called she will do the test as soon as possible and send it back in the courier bag. [Pasifika clinic] |
| Didn't understand what it was/poor English | 3 | Did not understand English so spoke to young son who translated. [Asian clinic] |
| Doesn't want GP or nurse to do the test | 3 | "I didn't keep the letter, I get embarrassed and don't want the nurse or doctor going there. If I can do it myself then I will try." [Pasifika clinic] |
| Never had sex | 3 | Has never had sex so didn't do the test and didn't think she needed to call. [Asian clinic] |
| Will do test herself | 3 | Yes, she knows about the study and yes she will do it when she gets around to it. [Māori clinic] |
| Will take test to GP | 2 | Still has the kit and information so will take with her to the doctor. [Asian home] |
| Will go for normal smear test | 2 | Participant lost the kit when moving house (before Christmas). Wasn't really interested as too busy. Will sort out doing her smear when she has more time. [Māori home] |
| Embarrassment/sacred body | 2 | "I received it however I take no interest in things like this, ... this is an invasive process not just self-sampling but smears as well and I was raised that this is a sacred part of my body and to be treated with respect. I have had smears in the past and was uncomfortable with the invasiveness of them. ... if I feel the need, I will seek what help I want." [Māori clinic] |
| Invasive procedure | 2 | Didn't like the idea of 'sticking anything up there'. [Asian home] |
| Will seek help if needed | 2 | "[-]I have had smears in the past and was uncomfortable with the invasiveness of them. I understand that this is just my view. I am not interested in any of these initiatives and if I feel the need I will seek what help I want. [-]" [Maori clinic] |
| Other responses (each N=1) | 8 |  |

Received all information and texts. She did not do the test as she does not want to do it. When I asked if there was anything that would have encouraged her to, she said no she just did not want to do it. [Pasifika home]

### Using the self-test kit

Forty-two women provided responses other than N/A or ‘I didn’t watch them’ to the open-ended question asking for comments about the video clips. The most frequent response was that they were clear and easy to understand (N=18). Eight women found the written or nurse instructions sufficient. Four women found them reassuring, four were too busy to watch them, two women were still uncertain about the test, two women commented on the different languages (one positive, one suggesting they could be more than just subtitles) and the rest of the responses mainly concerned technical issues (e.g., “Because video clips not available”).

### Barriers to smear test

Forty-one women provided a free text self-reported reason for not having had a smear test. These included dislike of test or fear of what might be found or needed as next steps (N=6), personal or relative’s previous bad experience of a smear test (N=6), shyness, embarrassment or modesty (N=6), too busy working, not bothered or living elsewhere (N=5), current or recent pregnancy (N=5), recently had the test or felt that the current schedule is too frequent (N=4), believing that they had not had exposure to HPV or didn’t need the test because they were a lesbian (N=3) and 6 miscellaneous reasons (e.g., looking for new doctor).

## Footnotes

1 Pasifika (sometimes spelt Pasefika) is used to refer to the people, cultures, and language of Pacific groups including: Sāmoa, Tonga, the Cook Islands, Niue, Tokelau, Tuvalu, and other smaller Pacific nations – who are now living in Aotearoa New Zealand. We acknowledge that there is “no generic ‘Pacific community’” and that the term Pasifika is “a category defined by New Zealand policy and discourse”10.

2 There is critique of the term whakamā in the interpretation of women’s reporting about the invasiveness of vaginal examinations/cytology test, with the use of these terms seen to be victim blaming/deficit framing. We deliberately refer here to the invasive nature of the test to reflect that it is the test that is problematic.

3 While both self-testing and self-sampling are used in the literature, self-testing was found to be the term preferred by women. Accordingly, that is the terminology we have used throughout the manuscript.

